# Short-Term Pediatric Acclimatization to Adaptive Hearing Aid Technology

**DOI:** 10.1101/2020.09.24.20100933

**Authors:** Joseph Pinkl, Erin K. Cash, Tommy C. Evans, Timothy Neijman, Jean W. Hamilton, Sarah D. Ferguson, Jasmin L. Martinez, Johanne Rumley, Lisa L. Hunter, David R. Moore, Hannah J. Stewart

## Abstract

**Purpose:** This pilot study assessed the perceptual, cognitive and academic learning effects of an adaptive integrated directionality and noise reduction hearing aid program in pediatric users.

**Methods:** Fifteen pediatric hearing aid users (6 to 12 years old) participated in a hearing aid pilot with pre- to post-comparisons. Participants received new bilateral, individually fitted Oticon OPN hearing aids programmed with OpenSound Navigator (OSN) processing. Word recognition in noise, sentence repetition in quiet, nonword repetition, vocabulary learning, selective attention, executive function, memory, reading and mathematical abilities were measured within one week of the initial hearing aid fitting and two months post-fit. Caregivers completed questionnaires assessing their child’s listening and communication abilities prior to study enrollment and after two months of using the study hearing aids.

**Results:** Caregiver reporting indicated significant improvements in speech and sound perception, spatial sound awareness and the ability to participate in conversations. However, there was no positive change in performance in any of the measured skills. Mathematical scores significantly declined after two months.

**Conclusions:** OSN provided a perceived improvement in functional benefit, compared to their previous hearing aids, as reported by caregivers. However, there was no positive change in listening skills, cognition and academic success after two months of using OSN. Findings may have been impacted by reporter bias, limited sample size and a relatively short trial period. This study took place during the summer when participants were out of school which may have influenced the decline in mathematical scores. The results support further exploration with age and audiogram-matched controls, larger sample sizes, and longer test-retest intervals that correspond to the academic school year.

## Introduction

Sensorineural hearing loss (SNHL) is an impairment that decreases hearing sensitivity and degrades suprathreshold sound perception (Plomp, 1978). This is a leading cause of perceptual, social and academic disability in children (Finitzo-Hieber & Tillman, 1978; Hick & Tharpe, 2002; Leibold et al., 2013; Levy-Shiff & Hoffman, 1985; Most et al., 2012). Hearing aids (HAs) provide sound amplification as an intervention for SNHL and offer individualized programming capabilities to optimize treatment on a case-by-case basis. For pediatric patients, programming strategies have historically favored omnidirectional microphones because spherical polar patterns detect sounds equally from all directions and were thought to promote incidental learning (American Academy of Audiology, 2004; Cunningham, 2007). While omnidirectional amplification may increase overall sound exposure, it does not mitigate the suprathreshold effects of SNHL that can impair speech perception, particularly when background noise is present (e.g. Killion, 1997; Leibold et al., 2013; Lesica, 2018). If sound perception is degraded, it negatively impacts further processing of speech, potentially diminishing the benefits of language exposure.

Interfering noise is a major challenge to HA treatment for SNHL because it impairs speech perception and cannot be completely isolated from the HA output (e.g. Chung, 2004 Park et al., 2015; Plomp, 1986). Compared to adults, this issue is greater for pediatric patients because children typically have poorer speech perception and spend more time in environments with interfering noise (Crukley et al., 2011). School environments in particular contain high noise levels, sometimes reaching 37 decibels (dB) below the recommended +15 to +30 dB signal-to-noise ratio (SNR; American National Standards Institute, 2002; Picard & Bradley, 2001). The masking effect of noise can obscure phonetic coding and auditory stream formations leading to reduced speech comprehension and for children, potential delays and inaccuracies in language learning (Hawley et al., 2004; Riley & McGregor, 2012; Shinn-Cunningham & Best, 2008). Degraded speech requires further cognitive effort to complete missing or distorted phonetic information, thereby reducing information intake, working memory capacity and selective attention (Hick & Tharpe, 2002; Pichora-Fuller et al., 2016; Rönnberg et al., 2013; Shinn-Cunningham & Best, 2008), making a child poorly primed for incidental and intentional learning experiences (Bregman, 1990; Gilbertson & Kamhi, 1995; McCreery & Stelmachowicz, 2013; Moore et al., 2008; Szalárdy et al., 2019). These disadvantages appear to diminish at favorable SNR levels, as Stiles and colleagues (2012) reported no significant difference in working memory and articulation rate in six to nine year olds with mild to moderately-severe SNHL when completing listening tasks at +15 dB SNR and in quiet. Finding ways to reduce noise interference in pediatric HA users is therefore warranted since speech intelligibility, working memory and selective attention all connect to multiple areas of academic success (Locke, 1997).

HA features such as digital noise reduction (DNR) and fixed-directional microphones can limit the output of unwanted noise, but their outcomes and practicality are not always optimal for children. Modulation-based DNR algorithms can improve perceptual sound comfort (Mueller et al., 2006; Ricketts & Hornsby, 2005) but do not increase within-channel SNRs, providing little benefit for speech intelligibility (Ahmadi et al., 2018; Crukley & Scollie, 2014; Peeters et al., 2009). Fixed-directional microphones are more effective in improving on-frequency SNR, with laboratory experiments showing a 2-5 dB SNR benefit (Gravel et al., 1999; Hornsby & Ricketts, 2007; Park et al., 2015). However, this improvement can only exist if the HA user is facing the target sound source in an environment of low reverberation (Gravel et al., 1999; Ricketts et al., 2007) - a scene not typical of a classroom (American Academy of Audiology, 2013). Due to these limitations, and the fact that children often attend to one person at a time (their teacher or parent), personal microphone systems are typically the primary recommendation for children. These are wireless microphones that can be placed near a talker. The input is transmitted wirelessly to the listener’s HA through radio signals or Bluetooth, negating acoustic diffusion and decay of a target voice. This can increase the SNR by approximately 15 dB depending on programming (Hawkins, 1984) but they are not always practical as many teaching styles use group learning and multiple forms of media (Fathman & Kessler, 1992; Gillies, 2008; Lacina, 2005; Shield et al., 2015). There are also drawbacks to using these devices outside of the classroom, specifically when multiple target talkers are present (e.g. cafeterias and playgrounds), and they are rarely advised for full-time use (American Academy of Audiology, 2011). Nevertheless, personal microphone systems remain the most recommended auditory intervention for learning barriers experienced by children with HAs (American Academy of Audiology, 2011, 2013).

The search for more effective and convenient strategies has led to the development of more complex signal processing systems that may better facilitate speech perception and language development in children. Adaptive directional microphones are among the most notable of these features and are implemented in advanced HA models. These systems automatically alter dual microphone signaling to steer polar pattern null points toward the estimated direction of unwanted noise to improve multidirectional SNR. Paired with an automatic switching program, some HAs alternate between omnidirectional and directional modes based on environmental noise levels to selectively increase SNR during appropriate listening situations. However, there are limitations to these systems, particularly when multiple noise sources are present, causing the acoustic scene analysis to break down (Chung & Zeng, 2009; Ricketts et al., 2017; Ricketts & Henry, 2002; Summers et al., 2008; Wolfe et al., 2017). Although not perfect, and not intended to completely replace personal microphones for classroom learning, evidence suggests that adaptive and automatic switching microphones are effective promoters of speech perception and incidental language learning compared to omni- and fixed-directionality and thus have been recommended for children as early as six months of age (e.g. Dillon et al., 2014).

Browning and colleagues (2019) investigated the immediate effects of a HA signal processing system called OpenSound Navigator (OSN) by Oticon in 5 to 14 year old HA users with SNHL ranging from the mild to severe degree. OSN is an adaptive integrated HA system that uses a two channel spatial-microphone noise estimator in series with a minimum-variance distortionless response (MVDR) beamformer and a modulation-based DNR program. The MVDR beamformer creates null points within the HA polar pattern that conform to the spatial noise estimation while the DNR reduces the output of steady-state residual noise that diffuse past the MVDR null points (for further details see Le Goff et al., 2016). This is intended to function as a highly efficient and stable system capable of enhancing speech signals at all directions while reducing non-speech, off-axis environmental noise. Using a free-field word recognition in noise test (Figure 1) Browning et al. measured its efficacy by presenting target speech either in front (0° azimuth) or to the left side (−60° azimuth) of a listener. Masking noise was either constant speech-shaped noise (SSN) or intelligible two-talker speech masking (TTM) positioned behind the participant at +135° and −135° azimuth. OSN-enabled amplification was compared to omnidirectional amplification immediately after the HA fit using a within-subjects study design. Significant improvements of word recognition in the presence of SSN at both 0° and −60° speech presentations were found when using OSN but there was no difference between amplification strategies when listening in TTM. The researchers concluded that OSN was not inferior to omnidirectional processing when listening in TTM. However, it did not differentiate competing speech from target speech and thus its benefits are dependent on the listening setting and the type of background noise.

**Figure 1:**
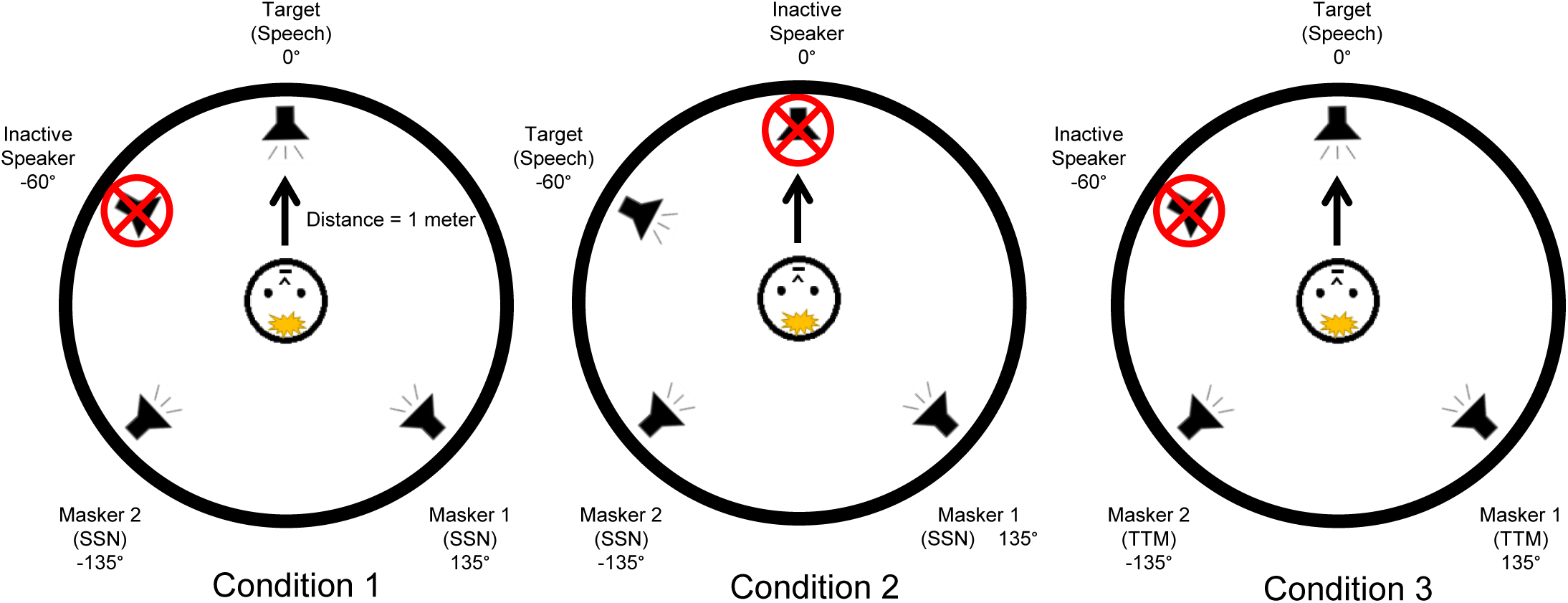
Words in noise test (Browning et al., 2019): Condition 1 involves speech presented in front of the listener at 0° azimuth with speech shaped noise (SSN) behind the listener at +/- 135°; Condition 2 involves speech presented to the side of the listener at 60° azimuth with SSN presented at +/-135°; and Condition 3 involves speech presented in front of the listener at 0° azimuth with two talker masker (TTM) presented behind the listener at +/-135°. All conditions were conducted in a 1m radius sound ring in a soundproof booth.

Browning et al. (2019) found no immediate benefit of OSN in TTM compared to omnidirectional amplification. However, through a form of perceptual learning called acclimatization, it is possible that users may learn to take advantage of newly available auditory cues to improve listening in noise skills and cognition (Arlinger et al., 1996; Gibson, 1969). Overall, evidence for HA acclimatization is mixed with some studies showing significant functional benefits after six to 16 weeks (e.g. Doherty & Desjardins, 2015; Horwitz & Turner, 1997; Munro & Lutman, 2003; Yund et al., 2006) and others showing minimal or no change in benefit within similar time spans (e.g. Dawes et al., 2014; Humes et al., 2002; Saunders & Cienkowski, 1997). Most of these studies however, focused on speech perception measurements in adults. Auditory plasticity is generally considered to be greater during early childhood (Boothroyd, 1997; Eggermont, 2008; Markman et al., 2011; Newport, 1990; Pulsifer et al., 2003; Su et al., 2008; Svirsky et al., 2000; White et a., 2013), potentially making children more sensitive to HA acclimatization and, by extension, cognitive and language development and academic learning (Willott, 1996). In this exploratory study we investigated the short-term effects of OSN on speech listening, language, memory, attention, executive functioning and academic performance in pediatric HA users over a two month period. We hypothesized that consistent use of OSN would promote acclimatization and further development in these skills. The methodology and findings of this study were used to develop our ongoing work involving a registered randomized controlled trial (RCT).

<Figure 1 about here - words in noise test>

## Methods

### Participants

Fifteen experienced pediatric HA users ages 6;4 to 12;8 (years;months, *M* = 9;10) were recruited through the audiology patient base at Cincinnati Children’s Hospital Medical Center (CCHMC; see Table 1). Parents and legal guardians were asked to complete a background questionnaire to help assess eligibility. Inclusion criteria included a) native English speakers, b) no history of ear surgeries, c) symmetrical SNHL in the mild to moderate-severe range in the frequency range of 500-4000 Hz (Sweitzer, 1977), d) no history of developmental delays, e) no medical diagnoses of neurologic/psychiatric disorders or attention deficits, f) history of consistent binaural HA use as reported by the child’s attending audiologist and medical notes, g) no prior experience with Oticon’s OSN algorithm and h) magnetic resonance imaging (MRI) compatible (to collect brain imaging data, to be reported elsewhere). Prior to testing, caregiver consent was obtained and documented. Assent was obtained from participants 11 years and older. Ethical approval for all clinical tests, services and data collection procedures were obtained from the CCHMC Institutional Review Board before initiating the study. All participants received financial compensation and were offered the opportunity to keep the study devices, earmolds and accessories free of charge upon study completion.

**Table 1:** Participant demographics and hearing aid history. Due to potential confidentiality breaches, details of participants’ ages have been removed from this copy of the paper.

| Participant ID | Age at Enrollment (years;months) | Sex | Maternal Education | Etiology | Age of Diagnosis (years;months) | Age at First HA Fit (years;months) | HA manufacture at Enrollment | HA Model at Enrollment | Time with Previous HA's at Enrollment (years;months) | Trial Period (time of research visit 1 to time of research visit 2; days) | Average Daily use of OSN During Trial Period (hours) |
| --- | --- | --- | --- | --- | --- | --- | --- | --- | --- | --- | --- |
| ot_001 |  | M | College degree | Unknown |  |  | Phonak | Sky Q-M 13 |  | 65 | 10.1625 |
| ot_002 |  | M | Some college | Enlarged Vestibular Aqueduct |  |  | Phonak | Bolero V70-P |  | 66 | 10.9 |
| ot_003 |  | F | College degree | Genetic |  |  | Phonak | Sky Q50 M13 |  | 72 | 10.95 |
| ot_005 |  | M | College degree | Genetic |  |  | Phonak | Sky Q50 M13 |  | 56 | 12 |
| ot_008 |  | M | College degree | Genetic |  |  | Oticon | Sensei Pro |  | 70 | 6.625 |
| ot_014 |  | M | College degree | Unknown |  |  | Oticon | Sensei Pro |  | 60 | 9.745 |
| ot_018 |  | F | Post-graduate degree | Unknown |  |  | Phonak | Sky Q70 M13 |  | 63 | 7.815 |
| ot_026* |  | F | College degree | Genetic |  |  | Oticon | Sensei Pro |  | 71 | 2.4525 |
| ot_027 |  | F | Post-graduate degree | Unknown |  |  | Oticon | Sensei Pro |  | 57 | 10.25 |
| ot_030 |  | M | High school | Unknown |  |  | Phonak | Sky V50-P |  | 66 | 11.5625 |
| ot_036 |  | F | No high school | Genetic |  |  | Oticon | Sensei Pro |  | 73 | 10.84 |
| ot_039 |  | M | Some college | Unknown |  |  | Oticon | Sensei Pro |  | 65 | 12.525 |
| ot_041 |  | M | College degree | Genetic |  |  | Oticon | Sensei Pro |  | 70 | 7.2 |
| ot_044 |  | F | Some college | Enlarged Vestibular Aqueduct |  |  | Phonak | Bolero V70-P |  | 73 | 12.45 |
| ot_048* |  | F | College degree | Unknown |  |  | Phonak | Sky V50-P |  | 67 | 9.5 |

**Figure 2:**
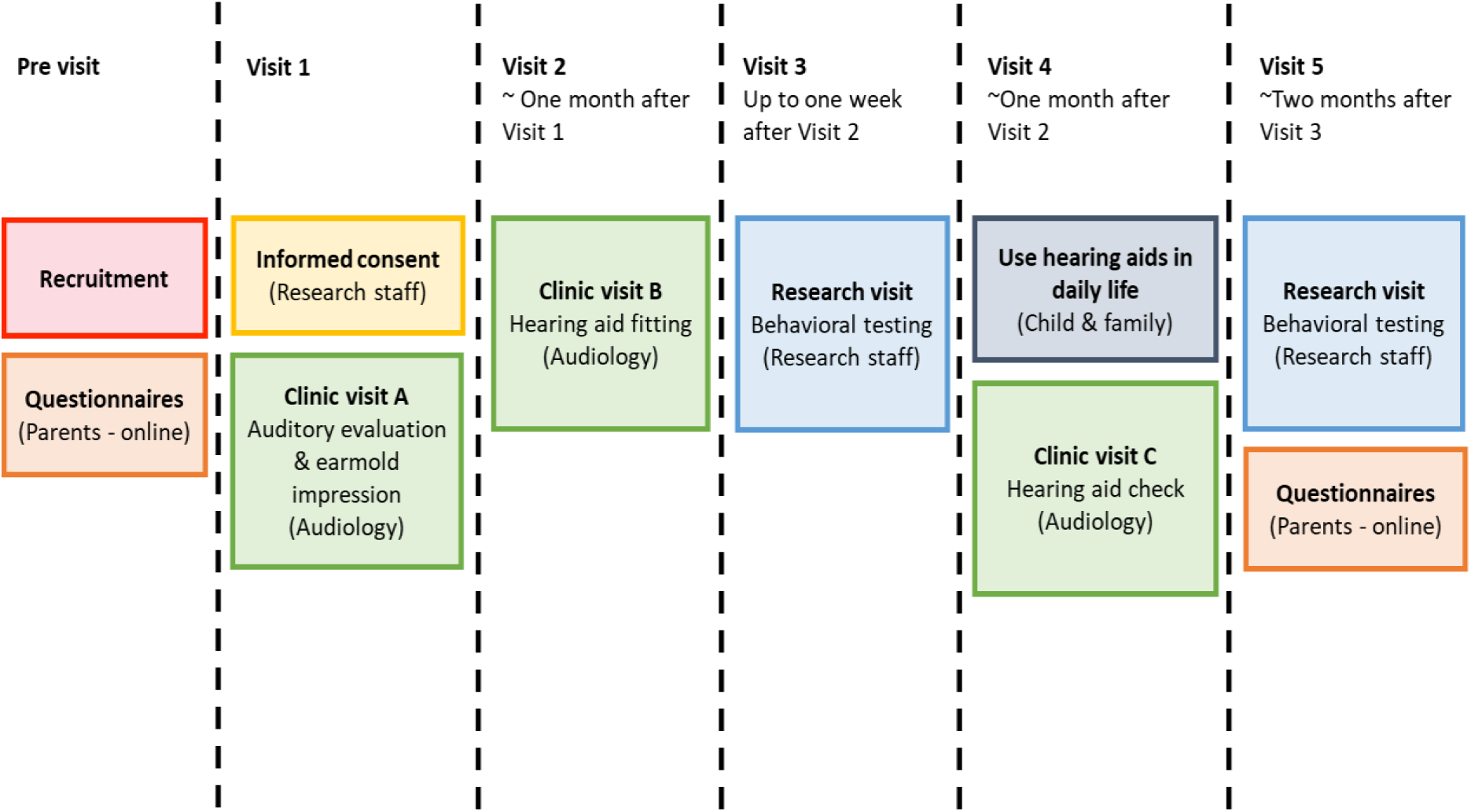
Detailed timeline of the study. Visit 3 involved data collecting at time point 1: T1 and Visit 5 involved data collecting at time point 2: T2.

### Protocol

The study followed a specific sequence of three clinic and two research visits over approximately three months (Figure 2). All clinic and research visits took place during the months of May to August, during the school summer holiday break.

### Audiometry

#### Audiologic evaluation

Clinical evaluation and HA fitting, programming and servicing were performed by licensed audiologists at the Division of Audiology at CCHMC. The evaluation included standard 226 Hz tympanometry, pure-tone thresholds from 250-8000 Hz at half-octave increments via air-conduction and 500-4000 Hz at octave increments via bone-conduction (see Figure 3 for air-conduction results), speech reception thresholds (SRT), and speech recognition in quiet. Tympanometry was completed using a GSI Tympstar Middle Ear Analyzer (Grason-Stadler, Eden Prairie, MN) or Titan/IMP440 (Interacoustics, Eden Prairie, MN). Air and bone-conducted signals were presented through EARTone 3A insert earphones and a MelMedtronics B71 adult bone oscillator, respectively using a GSI 61 Clinical Audiometer (Grason-Stadler, Eden Prairie, MN). SRTs were measured using a recorded male voice of closed set spondee words. Speech recognition testing was performed monaurally and binaurally using recorded male voices with open-set lists (NU-6, W-22 or PKB) through EARTone 3A inserts presented at 40 dB SL (sensation level; based on SRT) or the participant’s most-comfortable level. Previous audiometric results were used for participants who received an audiologic evaluation at CCHMC within six months of study enrollment. Ear mold impressions were taken at the audiologic evaluation.

**Figure 3:**
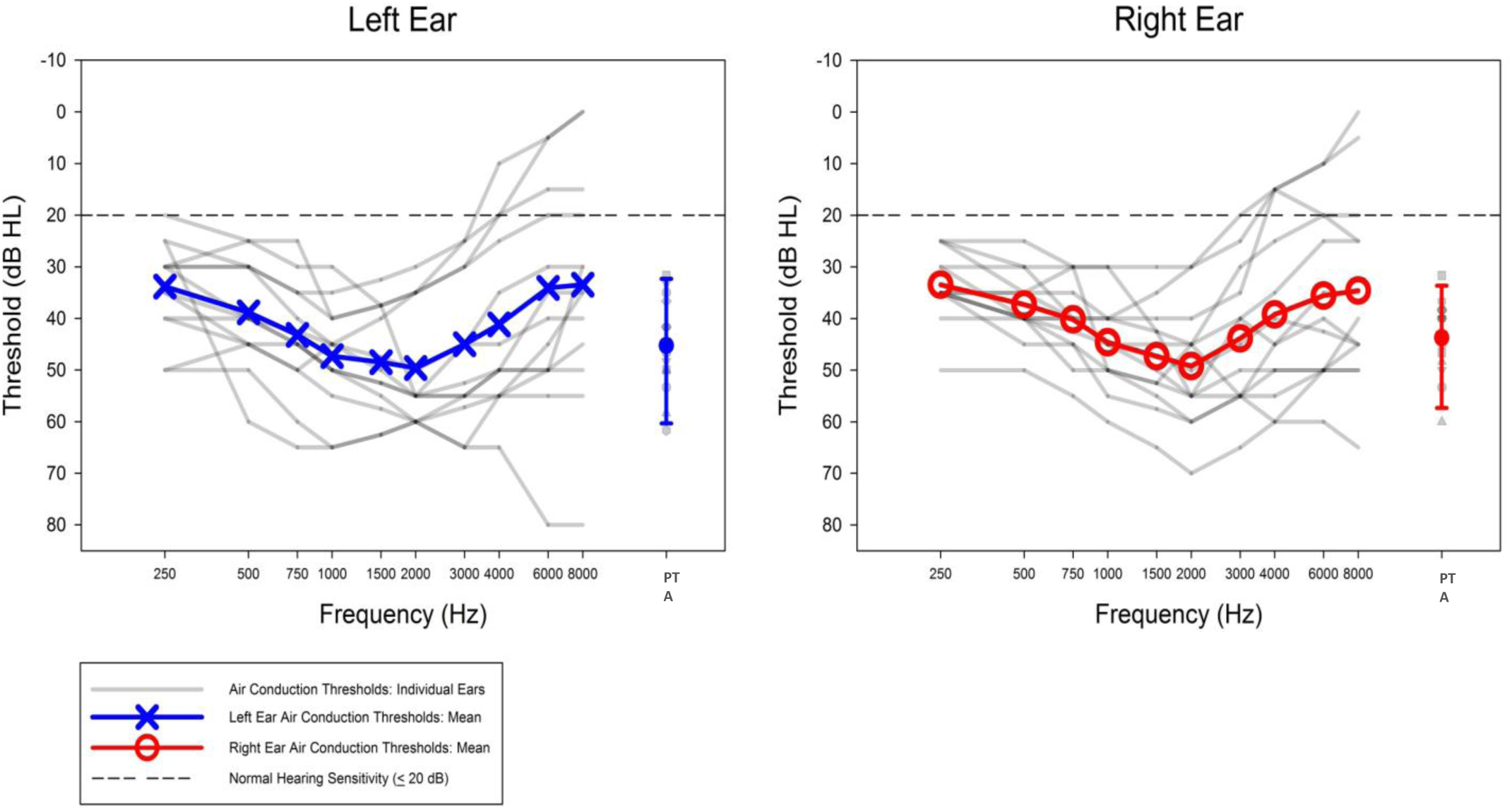
Line graphs for all audiograms are plotted (in gray) with a computed group average for the left ear (in blue) and right ear (in red). The pure tone averages (PTA) are calculated as the average of 500, 1000 and 2000 Hz thresholds, separately for each ear. PTA error bars indicate the 10th-90th percentiles.

### Hearing Aid Fitting

Within one month of the audiologic evaluation, bilateral Oticon OPN 1 PP behind-the-ear (BTE) HA’s with standard tubing, custom ear molds and a compatible wireless microphone and Bluetooth streamer, called ConnectClip were dispensed for each participant. Ear molds (Emtech laboratories, Roanoke, VA) were either skeleton or canal style and made from silicone material with venting size selected by the fitting audiologist. Manual volume control, manual program options, frequency lowering and feedback cancellation were disabled. The OSN algorithm was enabled with the transition and noise reduction in simple/complex environments set to the manufacturer default setting.

HA verifications were performed using Verifit 2.0 (Audioscan, Dorchester, ON, Canada). Real-ear speech mapping was completed using a standard recorded passage presented at 55, 65 and 75 dB sound pressure level (SPL). Prescriptive targets were set using Desired Sensation Level (DSL) v5.0 (Scollie et al., 2005) based on each participant’s audiometric thresholds and individual real ear-to-coupler differences. Real-ear probe microphone measurements were used to ensure that HA gain met prescriptive targets. Fine-tuning gain adjustments were made so that HA output was within 5 dB at 250, 500, 1000 and 2000 Hz and 8 dB at 3000 and 4000 Hz of the prescriptive targets (Bagatto et al., 2011; Bagatto et al., 2016; figure 4). Participants and accompanying parents/legal guardians were instructed on HA use and care. Participants were advised to use the ConnectClip for transmitting in their classroom setting.

HA follow-up appointments were completed at one month post fitting by the child’s fitting audiologist. Validation measures included narrow-band noise at center frequencies of 500, 1000, 2000 and 4000 Hz and speech recognition in quiet testing (NU-6, W-22 or PBK lists) using recorded male voices presented at 35 and 55 dB HL all of which were completed using a sound field at 0° azimuth in the binaurally aided condition. Word lists were consistent between the initial audiologic test and the follow-up testing. All participants reported satisfaction with the sound quality of the HAs at the time of the follow-up. Three participants reported minor physical discomfort and/or poor fit of the custom earmolds at the one month follow-up. Earmold remakes were dispensed within two weeks of the follow-up appointment. In the meantime, participants agreed to continue wearing the existing earmolds.

### Hearing aid usage

At the end of the study, average daily HA use was obtained from the data logging feature available on the fitting software (see Table 1).

**Figure 4:**
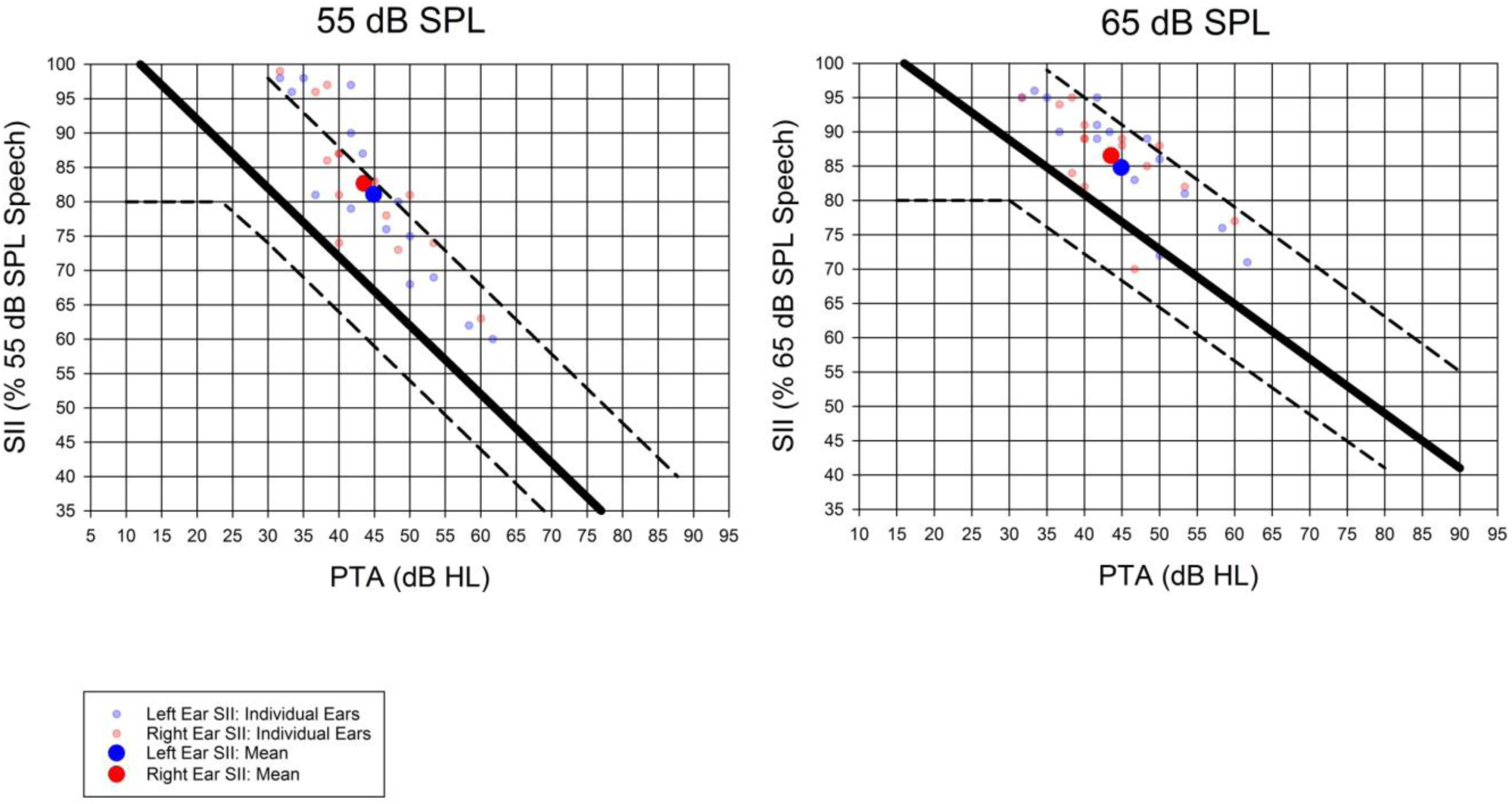
Speech intelligibility index (SII), a percentage value that estimates the intelligibility of speech of a patient based on audibility from 500-4000 Hz (American National Standards Institute, 2017), was calculated via aided real-ear aided measurements for soft speech (55 dB SPL) and moderate speech (65 dB SPL). Individual and mean SII values for the left right ears were plotted as a function of pure tone average (PTA) for 55 dB and 65 dB level of speech input. The solid line is the average SII for children from birth to age six. The lower dotted line represents one standard deviation below and the upper dotted line represents two standard deviations above the average SII. Best practice recommends aided SII’s to fall within or above the dashed lines (Bagatto et al., 2016).

### Experimental testing

All experimental behavioral tests were performed in a soundproof booth (IAC Acoustics, North Aurora, IL) with the participants wearing their Oticon OPN HAs and with OSN enabled. Unless otherwise stated, task stimuli were presented through an M-Track Eight interface (M-Audio, Cumberland, RI) Servo 120a power amplifiers (Samson Technologies, Hicksville, NY) and JBL Control 1Xtreme 4” loudspeakers (Harman International Industries, Stamford, CT) with the tester outside the test booth. For communication and response recording, the participant used a proximally positioned cardioid microphone while the tester used a headset with a wireless transmitter.

The order of testing was randomized in a latin square design across participants. All tests were completed twice: within one week of the HA fitting (time point 1; T1) and 2 months post fitting (time point 2; T2).

### Listening in noise

*Word repetition:* SNR thresholds were measured using the same software and stimuli as Browning et al. (2019). Three different listening conditions were used (Figure 1). Target words were produced by a native English speaking, monolingual male (F0 average of 102 Hz) and presented at a fixed intensity of 65 dB SPL with an adaptive noise level set at an initial level of 55 dB SPL. Each TTM speech stream in Condition 3 was a recording of two monolingual male talkers in Standard American English (F0 averages of 121 and 140 Hz) reading separate passages from the children’s book *Jack and the Beanstalk*. Two separate streams of SSN were created based on the spectral envelope of the TTM speech streams for Conditions 1 and 2.

Target speech and noise stimuli were presented in a free-field sound ring (radius 1m) through custom software presented on Max 7 (San Francisco, CA) supplied by Boys Town National Research Hospital. The participant was situated in the center of the sound ring facing 0 ° azimuth using a chin rest for consistent HA microphone positioning. Participants were instructed to keep their head still, listen closely to the target word and repeat the word back. If they were unsure of what word they heard, they were instructed to guess. If the tester did not hear the response clearly, the participant was asked to verbally spell out the word (misspelling did not affect scoring).

If the repeated word was correct, the noise level increased by 4 dB, if incorrect, the noise reduced by 4 dB. After the second reversal, the adaptive noise level changed to 2 dB increments. This continued for six more noise level reversals for an overall SNR threshold calculation. The final threshold score indicates the estimated SNR level in which the participant correctly recognizes monosyllabic words 50% of the time (i.e. SNR-50).

### Listening in quiet

*Sentence repetition:* To assess speech perception while controlling for verbal working memory and grammatical knowledge, participants listened to and repeated back recorded sentences, following the work of Moll et al. (2015). The procedure was further developed to create a second version, allowing testing at two time points. Sentences were categorized based on length (use or non-use of adjuncts or adjectives) and morphosyntactic complexity (use or nonuse of transitive, intransitive and/or ditransitive verbs). Twenty sentences were used in each task list, made up of five sentences of each of the four sentence categories: short with low complexity (SL; e.g. "The man served a woman the fruit.”); short with high complexity (SH; e.g. "A piano was delivered by the dad to his son.”); long with low complexity (LL; e.g. "A helpful boy saved the good girl a silver spoon.”); and long with high complexity (LH; e.g. "The crumpled magazine was passed by a pretty woman to the tall boy.”).

Sentences were presented as recordings of a native female speaker in Standard American English and presented at 50 dB SPL using an online research platform provided by Uppsala University *(Audio Research*, n.d.). Two loudspeakers were situated 84 cm in front of the listener. The participant was instructed to face the loudspeakers and listen closely to the voice and repeat back the sentence. If the participant forgot or did not hear a word, they were encouraged to guess. The tester scored each sentence based on the accuracy of word content and order. Each test session was recorded using Audacity (v. 2.3.2). After testing was completed the recording was re-scored by a different researcher. If there was agreement between the two researchers, the initial score was used as data. If there were any disagreements between the initial score and the second score, a third researcher scored the test. Scores that received agreement from two scorers were used for the analyses.

### Novel word learning

*Nonword repetition:* NEPSY-II (Brooks et al., 2009; Korkman et al., 2007a, 2007b) is a US standardized neuropsychological test battery for children ages 3 to 17 that measures multiple neurocognitive domains. The subtest Repetition of Nonsense Words assesses novel word learning, specifically the ability to encode and decode phonological stimuli and articulate novel words. Children were presented with 13 recorded nonsense words of a male’s voice in quiet and were tasked with repeating the words back. Each word has two to five syllables and one point was awarded for each correctly recited syllable. Points for all words were summed together for a composite score that was then converted to an age-adjusted scaled score. Words were presented at 65 dB SPL using the same loudspeaker configuration as the sentence repetition test in quiet. The recording was re-scored using the same procedures used for the sentence repetition test in quiet.

### Cognition

The NIH Toolbox® (NIH-TB) is a collection of standardized comprehensive and interactive behavioral tests that measure multiple neurocognitive domains (Weintraub et al., 2013) via an iPad (Apple, Cupertino, CA). The program offers multiple subtests that take five to 15 minutes each to complete. Each test yields a raw score to be converted into an age-corrected score. Four NIH-Cognition-TB subtests (below) were used in this study allowing an overall ‘early childhood composite score’ to be calculated. Participants completed the tasks while comfortably seated at a table with the tester seated next to them.

*Picture Vocabulary (PV)* measures accuracy of word identification and vocabulary knowledge. A randomized set of words were spoken by a female in Standard American English. During each word presentation, four different pictures were presented on the iPad screen. The participant was tasked with selecting the picture that matched the word. If they were unsure what the word meant, they were instructed to make a guess.

*Flanker Inhibitory Control and Attention Test (FICA)* measures inhibitory control and selective attention. Five arrows (for eight year olds and older) or fish cartoons (for children under eight years) were presented on the iPad screen, each one pointing to the right or left. Using their index finger of their dominant hand, the participant was tasked with pressing the button that corresponded to the direction of the center arrow/fish irrespective of the directions of the other arrow/fish. Between trials participants had to keep their index finger on a central dot in front of the iPad nicknamed ‘homebase’. Scoring is based on accuracy and response time. The participants were asked to respond as quickly and as accurately as they could.

*Dimensional Change Card Sort Test* (DCCS) measures executive function by changing the rules that pairs of cards should be sorted by. In each trial a female voice stated whether the sorting rule was to match ‘shape’ or ‘color’. The participant used their index finger on their dominant hand to select their response and to wait at ‘homebase’ between trials. Scoring is based on accuracy and response time. The participants were asked to respond as quickly and as accurately as they could.

*Picture Sequence Memory Test (PSMT)* measures episodic memory capacity. Illustrations are presented in a specific sequence of events with a female voice briefly describing the image (e.g. "At the park we…play on the swings…fly a kite…" etc.). After their description, each image is left on the screen in the order of events. At the end of the trial all of the images are ‘shuffled’ and the participant must drag the images back into their correct order. Different versions of the test were used for the T1 and T2 visits.

#### Academic achievement

The Woodcock-Johnson IV (WJ-IV) Tests of Achievement is a standardized test battery used to measure academic achievement in children (McGrew et al., 2014). Each subtest is designed so that the questions gradually get more difficult. Questions were scored following a basal and ceiling rule (see McGrew et al., 2014 for scoring format). This study included four WJ-IV subtests allowing general reading (Letter-Word Identification and Passage Comprehension) and mathematical (Applied Problems and Calculation) scores to be calculated. Participants completed the tasks while comfortably seated at a table with the tester seated next to them. Different versions of the test were administered for the T1 and T2 visits.

*Letter-Word Identification* measures word identification and pronunciation. Groups of words were presented on the computer screen and the participant was tasked with reading them out from top to bottom.

*Applied Problems* measures a child’s ability to solve math-based problems in the form of sentences and/or graphics. For the more difficult problems, participants were offered to use a pencil and paper to work out the problems.

*Passage Comprehension* measures reading comprehension skills. For younger children (six years old) Passage Comprehension begins by presenting three pictures, the tester says a word or phrase, and the participant must point to the proper picture. As the test advanced, participants were presented with a sentence with a word missing. The child was tasked with reading the sentence in silence and verbalizing the missing word. Over the course of the test, the sentences became lengthy passages with more complex contextual cues making it more difficult.

*Calculation* measures mathematical knowledge. Participants are tasked with answering mathematical problems on paper. Problems advance from simple numerical problems to geometric, logarithmic and calculus.

### Caregiver reports

Two questionnaires: the Glasgow Hearing Aid Benefit Profile (GHABP; Gatehouse, 1999) and the Speech, Spatial and Qualities of Hearing Scale (SSQ; Gatehouse & Noble, 2004) were completed by parents or legal guardians. We used versions adjusted for caregiver assessment of their children’s abilities (GHABP: Kubba et al., 2004; SSQ: Galvin & Noble, 2013). Identical questionnaires were completed at two visits separated by about 2 months: during the T1 and T2 testing visits (Figure 2). The questionnaires asked about the child’s experiences and abilities prior to study enrollment (T1 data) and with the research Oticon OPN HAs (T2 data).

*GHABP:* This questionnaire includes 24 questions assessing change in hearing disability, handicap, HA use, HA benefit, HA satisfaction and residual (aided) disability relative to the benefit of their previous HAs (i.e T1 comparing two previous HAs and T2 comparing most recent HA to OPN). Questions used a five point Likert scale that was converted to numeric values (100 = much better, 50 = a little better, 0 = no change, −50 = a little worse, −100 = much worse; Kubba et al., 2004). Responses were calculated as an average composite score, with higher scores indicating greater HA benefit.

*SSQ parental version:* This questionnaire contains 27 items pertaining to speech hearing, spatial hearing, qualities of hearing, and conversational uses of hearing using a sliding 100 point scale (0 = not at all, 100 = perfect; Galvin & Noble, 2013). Scores for each section were averaged for a potential score ranging from 0 to 100 with higher scores indicating greater HA benefit.

### Analysis

Study data were collected and managed using Research Electronic Data Capture (REDCap) tools hosted at Cincinnati Children’s Hospital Medical Center. Statistical inferences were made by comparing experimental testing performance and caregiver reporting at ‘time point 1’ (T1) and ‘time point 2’ (T2) using univariate paired t-tests and repeated measures analyses of variance (rmANOVA). Post hoc t-tests with Bonferroni adjustments for multiple comparisons were computed when rmANOVA results indicated significant interactions and/or main effects. All data analyses were computed using Jeffrey’s Amazing Statistics Program (JASP) 0.9.0.1 and an alpha level of .05.

## Results

Data logging on the fitting software revealed full time HA use (≥ 6 hours per day bilaterally) for all participants except one child [ot_026] who used the HAs for an average of two hours per day (Table 1). This participant was removed from the data analyses. One participant [ot_048] reported a long-standing malfunctioning HA at T2. A listening check revealed intermittency and distortion. This participant was removed from the data analyses because it was unclear when and how long the issues occurred during the two month trial period. Formal testing (Figure 5) showed no significant change in performance after an average of 65.8 days of OSN experience, as detailed below. One exception was a decline in Mathematics scores over the trial period.

### Experimental testing

#### Listening in noise

*Word repetition (Figure 5A):* Some initial, sporadic technical difficulties were experienced with the testing software throughout the study leading to inaccurate and missing SNR threshold calculations. These data points were removed from the analyses leaving 6 data points for Condition 1, 11 for Condition 2 and 6 for Condition 3. Due to the unbalanced sample sizes across the three testing conditions, paired t-tests were computed individually for each test condition. No significant change in SNR threshold was found for any condition (Condition 1: t(5) = .16, *p* = .87, *d* = −.028, Condition 2: t(10) = 2.03, *p* = .069, *d* = −.645 and Condition 3: t(5) = 1.14, *p* = .31, *d* = −.76).

#### Listening in quiet

*Sentence Repetition (Figure 5B-5C):* There was no significant difference in total word repetition score between the two time points as revealed by a paired t-test (Figure 5B; t(11) = .17, *p* = .87, *d* = −.049). Subscores (Figure 5B) also failed to change significantly for content words (t(11) = .23, *p* = .80, *d* = .077) and function words (t(11) = .44, *p* = .67, *d* = −.13).

The effects of time, sentence length (short vs. long) and sentence complexity (low vs. high; Figure 5C) were investigated using a 2 × 2 × 2 rmANOVA. There were significant main effects for sentence complexity and length. Their interaction was also significant, as shown with post hoc t-tests revealing higher sentence repetition scores for short, low complexity (SL) sentences compared with all other sentence types, and for short, high complexity (SH) sentences compared with long, low and high complexity (LL and LH, respectively) sentences. There was no significant main effect of time, nor were there significant interactions with time (see Table 2A-2B for statistics).

**Table 2:** (A) Repeated measures analysis of variance of sentence repetition in quiet. Within subject factors of sentence length (short vs long), sentence complexity (low vs high) and time point (T1 vs T2) were used in the analysis. (B) Post hoc t tests with Bonferroni adjustments paired t-test of sentence repetition in quiet. The p values of significant main effects, interactions and difference of means are in bold. Note: SL= short sentences of low complexity, SH = short sentences of high complexity, LL = long sentences of low complexity, LH = long sentences of high complexity

| **A** |  |  |  |  |
| --- | --- | --- | --- | --- |
| **Source of Variation** | **df** | **F** | **p** | **$\eta^2$** |
| Length | 11 | 30.45 | **< .001** | 0.74 |
| Complexity | 11 | 15.62 | **0.002** | 0.59 |
| Time | 11 | 0.092 | 0.77 | 0.008 |
| Length * Complexity | 11 | 5.08 | **0.046** | 0.32 |
| Length * Time | 11 | 0.61 | 0.45 | 0.052 |
| Complexity * Time | 11 | 0.19 | 0.67 | 0.017 |
| Length * Complexity * Time | 11 | 0.014 | 0.91 | 0.001 |

**Table 2:** (A) Repeated measures analysis of variance of sentence repetition in quiet.
| **B** |  |  |  |
| --- | --- | --- | --- |
| **Comparison** | **t** | **p** | **Cohen's d** |
| SL vs. LH | 7.445 | **<0.001** | 1.73 |
| LL | 6.414 | **<0.001** | 1.52 |
| SH | 3.455 | **0.009** | 1.28 |
| SH vs. LH | 3.989 | **0.002** | 0.91 |
| LL | 2.959 | **0.034** | 0.68 |
| LL vs. LH | 1.031 | 1 | 0.19 |

One participant at T2 received an incorrect test version. Data from both testing sessions for this individual were removed, leaving 12 participants for the analyses.

#### Novel word learning (Figure 5D)

Mean scaled scores for NEPSY II Repetition of Nonsense Words were within the expected normative range of eight to 12 at both time points. There was no significant change in performance across trial time points (t(12) = 1.48, *p* = .17, *d* = −.41). Follow-up exploratory analyses showed a decreasing trend in variance (t(12) = 2.14, *p* = .054, *d* = .59) at T2 compared to T1.

#### Cognition (Figure 5E)

Mean NIH-TB age scaled scores were within one standard deviation of the age-adjusted normative score of 100 for all four subtests (PV, FICA, DCCS and PSMT). Using a 2 × 4 rmANOVA, results showed a significant difference between NIH-TB subtest scores (F(3, 36) = 5.60, *p* = .003, η^2^ = 32), but no main effect of trial period (F(1, 12) = .27, *p* = .64, η^2^ = 019) or interaction between time and subtest (F(3, 36) = .15, *p* = .93, η^2^ = .012). Post hoc t-tests revealed that scores were significantly higher for the PSMT subtest compared to the FICA (t(12) = 3.19, *p* = .039, *d* = .89) and DCCS (t(12) = 3.37, *p* = .033, *d* = .94). All other comparisons were nonsignificant (p ≥ .098).

The total age scaled Early Childhood Composite Score did not differ between the two time points (t(12) = .50, *p* = .62, *d* = .14).

#### Academic achievement (Figure 5F)

Woodcock-Johnson standard reading score (calculated from the Letter-Word Identification and Passage Comprehension subtests) did not differ significantly between time points (t(12) = .23, *p* = .82, *d* = .064). However, the mathematics score (calculated from the Applied Problems and Calculation subtests) did undergo a significant decline from T1 to T2 (t(12) = 3.25, *p* = .007, *d* = −.90).

**Figure 5:**
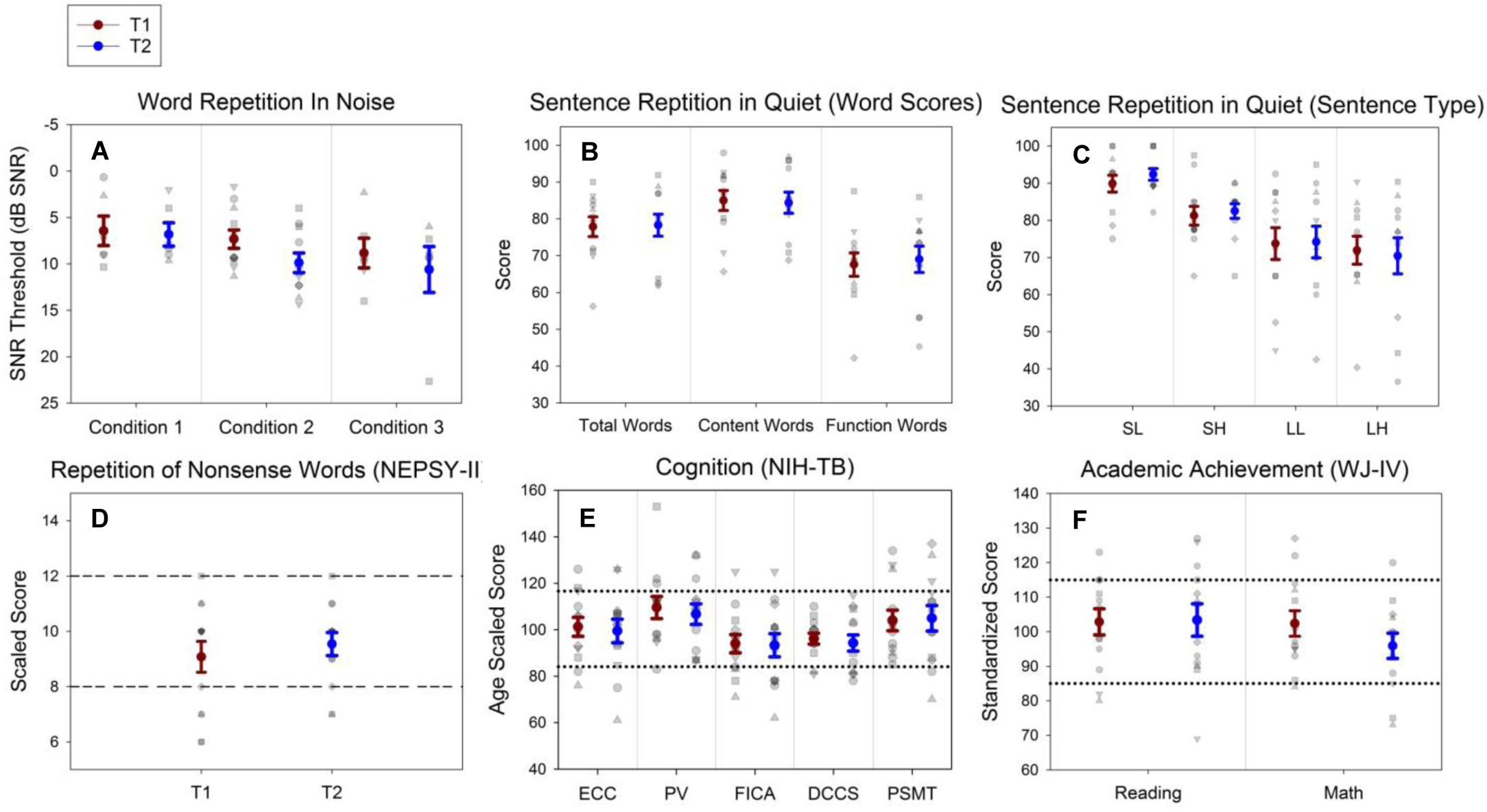
(A) SNR thresholds for Words in Noise for Conditions 1 (Target 0° azimuth and SSN), 2 (Target −60° and SSN) and 3 (Target 0° and TTM); sentence repetition scores in quiet for (B) total words, content words and function words and for (C) four sentence types: SL (short and low complexity), SH (short and high complexity), LL (long and low complexity) and LH (long and high complexity); (D) scaled scores for Repetition of Nonsense Words from NEPSY-II; (E) NIH-TB age scaled scores for ECC (Early Childhood Composite Scores), PV (Picture Vocabulary), FICA (Flanker Inhibition Control and Attention test), DCCS (Dimensional Change Card Sort), and PSMT (Picture Sequence Memory Test); (F) Woodcock Johnson-IV standard score for reading and mathematics. For all figures, the gray shapes indicate individual scores whereas the dark red and blue colored circles represent the mean score with error bars spanning +/- one standard error of the mean. For speech reception in noise testing, lower SNR thresholds indicate better performance. For all other tests, higher scores indicate better performance. For standardized tests, the normative range is indicated by the dashed line (NEPSY-II) and dotted lines representing one standard deviation above and below the normative score of 100 (NIH-TB and WJ-IV).

#### Caregiver reports

*Glasgow Hearing Aid Benefit Profile (Figure 6A):* No significant change in caregiver reported score was found between T1 and T2 time points using a paired t-test (t(12) = 1.48, *p* = .16, *d* = - .42).

*Speech, Spatial and Qualities of Hearing Parent Rating Scale (Figure 6B):* There was a significant main effect of time (F(1,12) = 10.33, *p* = .007, η^2^ = 46) reflecting higher reported overall scores at T2. There were no significant differences between subsection scores (F(3, 36) = 2.78, *p* = .055, η^2^ = 19) nor a significant interaction between subsection and period (F(3, 36) = 2.47, *p* = .078, η^2^ = .17), using a 2 × 4 rmANOVA.

**Figure 6:**
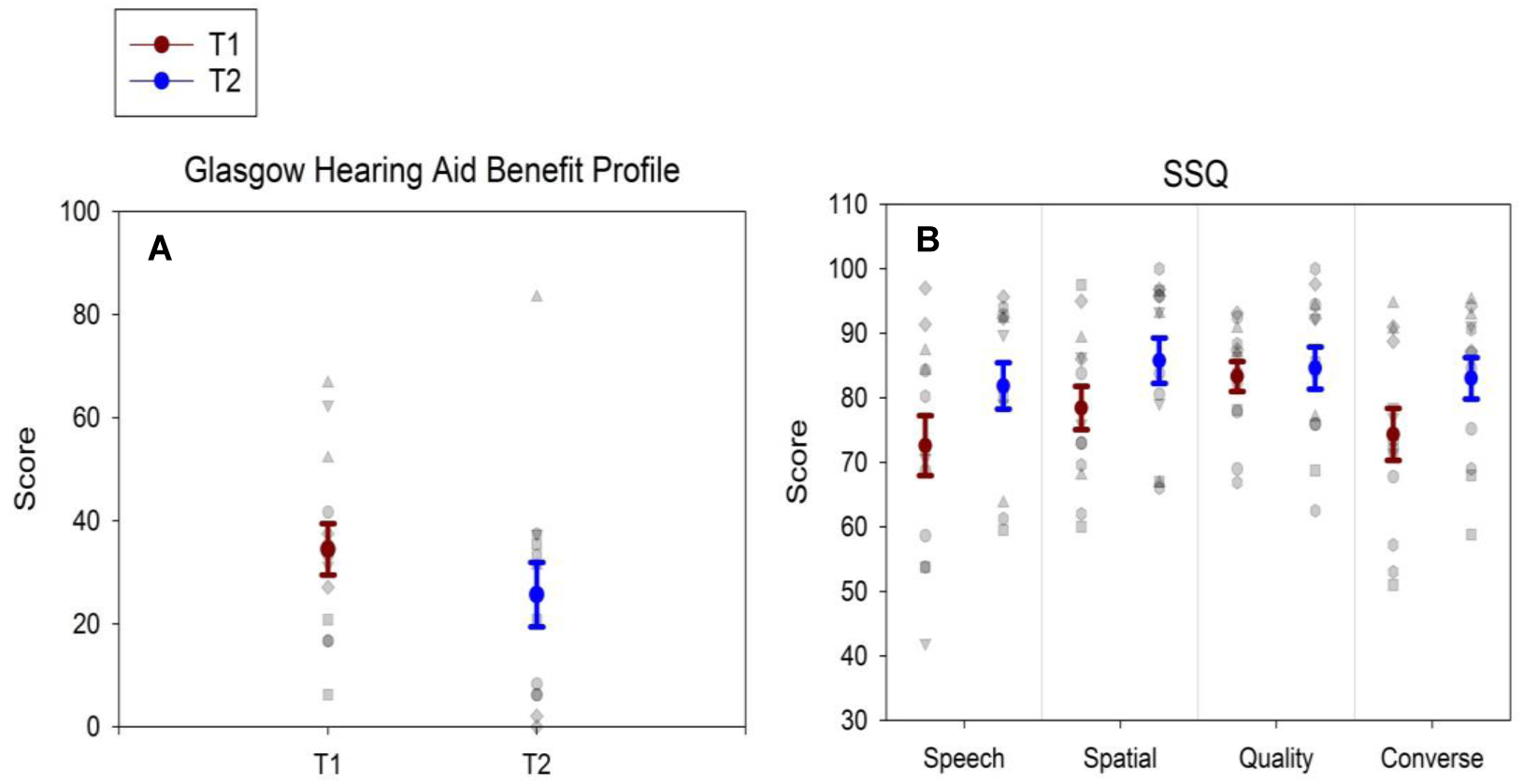
(A) Glasgow Hearing Aid Benefit Profile and (B) Speech, Spatial and Qualities of Hearing. The gray and white markings indicate individual scores whereas the brown and blue colored circles represents the mean score with error bars spanning +/- one standard error of the mean. On both questionnaires higher scores indicate better reported benefit.

## Discussion

OSN is intended to provide clear speech signals and spatial cues while reducing non-speech noise interference (Beck & Le Goff, 2017). We predicted that after two months of OSN use, children would improve listening skills, increase their learning rate of vocabulary and language, access newly available cognitive resources and improve academic performance. Compared to their child’s previous HAs, parents reported improved functional listening while using OSN. However, we found no evidence of improved speech perception, language development, cognition or academic achievement after two months of OSN experience.

### Language Learning and Processing

Degraded speech perception poses challenges for the speech processing pathway, potentially diminishing selective listening and language processing (Rönnberg, 2003; Rönnberg et al., 2013). Following OSN experience, we predicted improvements in speech repetition in noise but observed no such benefit for SSN or TTM. The masking effects of SSN and TTM are believed to reflect different mechanisms. SSN largely generates an energetic masking effect in which a spectro-temporal overlap obscures the perceptual resolution of the target stimuli, affecting auditory stream formations (Bregman, 1990; Moore et al., 2008; Szalárdy et al., 2019). In contrast, the masking effect of intelligible speech is in part informational-based, in which a failure of auditory stream selection occurs at what is believed to be a later stage of processing (Leek et al., 1991; Pollack, 1975; Srinivasan & Wang, 2008). Speech stream formation and selection abilities have been shown to have plastic properties and potential for training (e.g. Song et al., 2012) however our observations do not support acclimatized listening in noise abilities in pediatric HA users over the two month trial period.

In quiet, we found that sentence complexity and length influenced overall repetition performance, but we did not observe a significant change with OSN experience in sentence repetition ability. While this test is not standardized, Moll et al.’s (2015) study was well-powered and compared to their control data of typically developing six to 12 year olds (n = 57), our participants showed greater variation in their performance and, on average, scored lower for each of the four sentence types. Our findings were more similar to their experimental group of children with dyslexia (n = 40).

Evidence suggests that children with SNHL have poorer vocabulary, grammar and phonological processing skills compared to children with normal hearing, even when aided with HAs (Pittman, 2008; Pittman et al., 2005; Stelmachowicz et al., 2004). Consistent with this, we found that the children performed with lower accuracy on complex sentences compared to simple sentences suggesting difficulties with grammar. We also found that the children performed with lower accuracy on long sentences compared to short sentences, an effect that was widely believed to indicate impaired short-term and working verbal memory (Shankweiler & Crain, 1986). Previous evidence supports short-term memory difficulties in SNHL but primarily in novel word learning, nonword repetition, sequential short-term memory and competing noise tasks where long-term vocabulary and language knowledge have little influence on performance (e.g. Briscoe et al., 2001; DiGiovanni et al., 2017; Gilbertson & Kamhi, 1995; Hansson et al., 2004). It is possible that the effect of sentence length in this task reflects long-term language knowledge, receptive vocabulary and phonetic processing speed more so than short-term memory (MacDonald & Christiansen, 2002; Moll et al., 2015).

Nonword repetition was measured using NEPSY age-adjusted scores as an indicator of phonological processing ability and short-term phonetic memory (Coady & Evans, 2008; Moll et al., 2015). OSN was predicted to reduce the effort of listening that, over time, may improve phonological processing, working memory and short-term memory. While we found no improvement in NEPSY age-adjusted scaled scores at the group level, the mean age scaled scores were within the normative range of 8-12 at both time points. The variation in this measure reduced after two months of OSN use, however this reduction was not significant.

### Non auditory functions

Clearer speech signals and reduced noise levels enforce instructional learning and reduce the effort and fatigue of listening, possibly increasing the potential for other cognitive processes (DiGiovanni et al., 2017; Hornsby, 2013; Rönnberg, 2003). Using the NIH-TB to assess a range of cognitive skills we found that most children were within one standard deviation of the age scaled norms at T1 and T2 time points. We did not find a significant change in vocabulary, attention, memory and executive functioning skills after two months. Mean age scaled scores for picture vocabulary (PV) were the highest among all NIH-TB subtests and above the 100 point normative score at both time points. This may appear to contradict the sentence repetition findings which suggest language and vocabulary difficulties; however, the NIH-TB is a visual based vocabulary test (matching a picture to a spoken word) with no short-term memory demands unlike sentence repetition, which requires speech segmentation, encoding of grammar, articulation planning and speech production.

After two months of OSN use we found no significant improvements in academic reading or mathematical abilities (as measured by the WJ-IV). Unexpectedly, we found a decline in mathematical ability. This may be related to "summer learning loss”, a phenomenon that describes significant declines in achievement levels at the starting school year compared to the end of the prior school year (Cooper et al., 1996). Evidence suggests that this mostly involves mathematical skills (Atteberry & McEachin, 2016). The result highlights this study’s relatively short duration that took place during the long US school summer vacation. Improvement of this methodology would include randomized intervention and testing within the academic school year.

### Caregiver report

The GHABP, which was used to evaluate hearing disability and handicap along with HA use, benefit and satisfaction, decreased in total score by approximately 5% at the T2 visit. While this was not significant, it was unexpected. SSQ mean scores, on-the-other-hand, significantly improved, suggesting that caregivers observed a benefit of OSN, compared to their child’s previous HA, in real world situations including conversing, ignoring transient and sustained background noise, sound localization and differentiating voices and environmental sounds.

These two questionnaires approached HA benefit from different angles. The SSQ used a sliding numeric scale from 0-100 to evaluate functional and behavioral-based observations in different listening scenarios. The GHABP used a 5-point Likert scale to assess change in perceived benefit between a patient’s current HA compared to their previous one. Four participants used only one set of HAs prior to study enrollment, meaning that caregivers did not have a previous HA to compare when completing the questionnaire at the T1 visit. The perceptual change of a first time fitting is expected to be greater than an upgrade in HA technology for an experienced user (Dawes & Munro, 2017; Olson et al., 2013; Reber & Kompis, 2005) and may have skewed GHABP scoring.

### Limitations and Future Directions

There are four main limitations to this study. First, as mentioned above, this study was completed during the summer and participants had limited, if any, opportunity to use the HAs in an academic setting. This may have influenced our findings as the children may have had reduced multi-talker contact over the summer holidays and so had less opportunity to take advantage of the auditory cues provided by OSN. Furthermore, academic learning is an important contributor to vocabulary and language development as it provides direct instruction of word learning and grammar and facilitates the use of newly acquired vocabulary (Bowers et al., 2010; McKeown et al., 1985; Pellicer-Sanchez & Schmitt, 2010). Robust changes in vocabulary size and reading abilities may have been observed if our measurements were completed during the school year.

Second, we had intermittent software problems throughout the word repetition in noise testing. While Browning et al. (2019) found that children experienced an immediate OSN benefit when listening to a target in SSN, we did not find improvements after two months. However, our software issues reduced our data size for that test and may have increased the possibility for type 2 error due to a lack of power. Third, the comprehensive test battery included tasks of focused listening, problem solving, passage reading and various forms of memory recall that took about two hours to complete. To reduce the effect of fatigue the tasks were counterbalanced between participants and breaks were offered throughout the testing sessions. However, it was difficult to engage some of the younger participants. Lastly, caregiver reporter bias may have positively skewed the T2 SSQ mean score (e.g. Coughlin, 1990; Houtveen & Oei, 2007). We attempted to minimize re-test bias by prohibiting caregivers from seeing their initial responses when completing the questions at T2. It should be noted that we did not observe a ceiling effect.

This study was conducted as an exploratory study and while our findings do not support short-term acclimatization and learning effects in children, it is possible that a longer duration of OSN experience may yield positive outcomes. Perceptual learning, selective attention and executive function require functional changes within the central nervous system and without active training, may take longer than two months to acquire (Horwitz & Turner, 1997; Munro & Lutman, 2003; Ng et al., 2014; Ng & Rönnberg, 2020; Pereira-Jorge et al., 2018; Rählmann et al., 2018; Reber & Kompis, 2005). Furthermore, the lack of a control group prevents the ability to assess practice effects. Continuation with this methodology with an extended timescale may better characterize acclimatization effects in pediatric HA users.

Evidence for HA acclimatization has been mixed, with a paucity of pediatric-focused research. A potential reason for this is intersubject variability caused by numerous factors including age, degree and configuration of hearing loss, pre-lingual vs post-lingual onset, age of initial HA fit, etiology of hearing loss, and HA programming (e.g. compression ratios and knee points, and frequency nonlinearity). Our inclusion criteria and HA fitting protocol helped control for some intersubject variability, however, the study sample included participants with varying amounts of life-time HA experience and PTAs ranging from approximately 30-60 dB HL. In addition, potential for acclimatization may vary across our 6 to 12 year-old age group as perceptual sensitivity to speech and selective attention have been shown to develop at different rates from birth into adolescence (Halliday et al., 2008; Moore et al., 2008; Nittrouer & Lowenstein, 2007). Consequently, it is possible that intersubject variability obscured small but existing learning effects.

A more rigorous methodology to control for intersubject variability would be to assess two groups of age- and audiogram-matched listeners where each group is given a different HA algorithm. We have extended into such a trial with a double-blind RCT comparing long-term (at least six months) use of OSN to omnidirectional amplification. Results from this study were used to power this RCT. We intend to further our understanding of acclimatized listening in children and its effects on language development, cognition and academic performance when using adaptive HA technology.

## Data Availability

Data is only available to the authors

## Acknowledgments

The authors would like to thank Cecilia Nakeva von Mentzer for providing the sentence repetition task along with task interpretation, Boys Town National Research Hospital for supplying the word repetition in noise test software, Li Lin for her assistance with statistical analyses, and Elaine Ng and Thomas Behrens from Oticon for help with funding, providing hearing aids, and other general support. Thanks also to Anne-Marie Wollet, Jill Huizenga and Erin Stewart for help with some of the audiology visits.

